# A Blood-Based Biological Age Model Derived from Routine Laboratory Biomarkers in the Singapore Longitudinal Ageing Study

**DOI:** 10.64898/2026.05.05.26352428

**Authors:** Ye Jiangfeng, Xinru Lim, Maximilien Franck, Li Wenrui, Denise Goh, Chen Hao, Zara Chung, Wu Yang, Zhu Zhu, Patrick Henry Sitjar, Timothy Obi Wang, Felicia Wee, Rachel Elizabeth Ann Fincham, Daniel Shao Weng Tan, Kenon Chua, Tang Hong-Wen, Goh Jor Ming, Lim Wan-Teck Darren, Elaine Lim, Alan A Cohen, Roger Ho Chun Man, Tan Min-Han, Jia Gengjie, Tamàs Fülöp, Joe Yeong

**Affiliations:** Institute for Health Innovation and Technology (iHealthtech), National University of Singapore, Singapore; Institute of Molecular and Cell Biology (IMCB), Agency for Science, Technology and Research (A*STAR), Republic of Singapore; Research Center on Aging, Faculty of Medicine and Health Sciences, University of Sherbrooke, Sherbrooke, Quebec, Canada; Genome Analysis Laboratory of the Ministry of Agriculture and Rural Affairs, Agricultural Genomics Institute at Shenzhen, Chinese Academy of Agricultural Sciences, Shenzhen, China; Lucence Diagnostics Pte. Ltd., Singapore; Department of Biobank, The first affiliated hospital of Zhengzhou University, Zhengzhou, China; Department of Anatomical Pathology, Singapore General Hospital, Singapore; National Cancer Centre Singapore, Singapore; Duke-NUS Medical School, Singapore; Robert N. Butler Columbia Aging Center, Mailman School of Public Health, Columbia University, New York, USA; Department of Environmental Health Sciences, Mailman School of Public Health, Columbia University, New York, USA; The First Affiliated Hospital of Wenzhou Medical University, Wenzhou, China; Zhejiang Key Laboratory of Intelligent Cancer Biomarker Discovery and Translation, The First Affiliated Hospital of Wenzhou Medical University, Wenzhou, China; Department of Microbiology, Singapore General Hospital, Singapore

**Keywords:** Laboratory biomarkers, mortality prediction, risk stratification, risk factors, clonal hematopoiesis

## Abstract

Risk assessment in clinical practice depends largely on clinical phenotypes, including age, sex, body mass index, blood pressure and comorbidities. Routine laboratory data remain underutilised despite their accessibility and low cost. Using data from the Singapore Longitudinal Ageing Studies (n = 5,409; follow-up median 11.4 years), we developed a mortality prediction model based on routine laboratory biomarkers. We derived a biological age (age quotient, or AQ) score, and investigated its role as a mediator between lifestyle risk factors and mortality. Both models and association analyses were validated in the US National Health and Nutrition Examination Survey (n = 6,593) and UK Biobank (n = 290,949) cohorts. AQ was significantly elevated in deceased individuals (P<0.0001). AQ acceleration was also observed (P<0.0001). In overall survival discrimination, AQ outperformed chronological age (C-index 0.629 [SE 0.011] vs 0.606 [SE 0.011]), indicating superior prognostic prediction. Additionally, incorporation of AQ into a baseline model containing chronological age resulted in an improvement in model fit (likelihood ratio test, P<0.0001), consistent with incremental predictive value for mortality beyond chronological age alone. Mediation analysis supports a partial mediating role for AQ in the relationship between lifestyle factors and mortality. In a 57-patient subset, higher AQ was associated with increased *TET2* clonal hematopoiesis burden (β≈0.016 per +1 AQ year), suggesting a potential link between AQ acceleration, CH risk and diseases of aging, requiring validation in larger cohorts. We identified differential associations between lifestyle factors and groups of biological age components, indicating selective effects across biological systems. These findings provide an evidence-based framework for earlier and more accurate identification of high-risk individuals, offering a practical and easy-to-implement tool to inform preventive strategies.

## Main

Many predictive tools for mortality are developed within the context of specific disease, where they are derived from a pre-defined patient cohort and applied after clinical diagnosis ^1–6^. This approach largely focuses on outcome after illness, rather than identifying individuals at elevated risk before disease manifests, when preventive interventions are possible. Considering that global aging represents a major public health burden and it is often accompanied with an increased prevalence of chronic disease and multi-morbidity ^7^, early identification of individuals at high risk of mortality could enable targeted intervention to support healthier aging. However, current risk stratification approaches remain limited as it often relies on demographic and lifestyle variables such as age, body mass index (BMI) ^4^, alcohol consumption ^5^, and smoking status ^6^, which are known risk factors associated with increased mortality in the general population. While there are aggregate scoring systems that combine various variables, it oversimplifies complex biology. Subjective health assessments, including self-reported data ^8^ and clinical judgement ^9^, could be useful but may vary between individuals. Collectively, these approaches are generalised and unable to capture early or subtle physiological changes that precede clinical deterioration.

Blood tests form the backbone of population health screening and represent a minimally invasive, accessible, and cost-effective tool in clinical practice to capture haematological, inflammatory, and metabolic parameters. As a readily obtainable source of systemic health information, it offers a promising avenue for early detection of physiological changes associated with disease risk ^10^. However, the deeper prognostic potential of routine laboratory biomarkers remains largely underutilised as these biomarkers are often interpreted in isolation rather than in combination. Information contained within routine laboratory biomarkers could ultimately serve as a simple, objective, and scalable approach to identifying high-risk individuals, while also capturing the downstream biological effects of lifestyle habits.

Here, we studied 5,409 individuals of the Singapore Longitudinal Ageing Studies (SLAS) with a median 11.4 years of follow-up [interquartile range (IQR), 9.0-15.4 years; baseline data presented in **Table 1**], to develop a predictive model using routine laboratory biomarkers to stratify 5- and 10-year mortality risk. Participants were stratified into low-, middle-, and high-risk groups according to risk score tertiles, and Kaplan–Meier survival curves showed significant differences in 10-year survival among the groups (**Fig. 1a**). After assessing and reducing collinearity among 35 candidate laboratory variables, we developed a Cox proportional hazards model and derived corresponding 10-year mortality risk estimates. The final model included 9 variables: age, sex, neutrophil count, monocyte count, basophil count, red blood cell count, serum creatinine, serum albumin, serum and fasting glucose. This achieved concordance indices (C-index) of 0.629 in SLAS and 0.721 in NHANES, and 0.580 and 0.647, respectively, among individuals with biomarkers within the normal range (**Extended table 1**). Thereafter, we derived a risk scoring system (age quotient, or AQ) to estimate the biological age of SLAS individuals (**Fig. 1b**) as a predictor of mortality. Compared with deceased individuals within 10 years, survivors had a younger AQ relative to chronological age, and lower AQ acceleration (−1.07 [−6.44 to 5.17] vs. 3.65 [−3.46 to 13.17] and −1.47 [−6.88 to 4.45] vs. 1.24 [−5.02 to 9.63]), respectively (both P < 0.0001; **Fig. 1c-d**). Cox regression analysis demonstrated robust predictive performance of AQ for all-cause and cause-specific mortality including cancer, cardiovascular disease, respiratory disease, and diabetes mortality (**Table 2**). AQ association with all-cause and cause-specific mortality was also separately validated in the National Health and Nutrition Examination Survey (NHANES, n = 6,593) and UK Biobank (UKB, n = 290,949) cohorts, achieving a concordance index of 0.721 and 0.706, respectively, for all-cause mortality. Adding AQ to all models that included chronological age resulted in improvements in risk classification.

**Figure 1.**
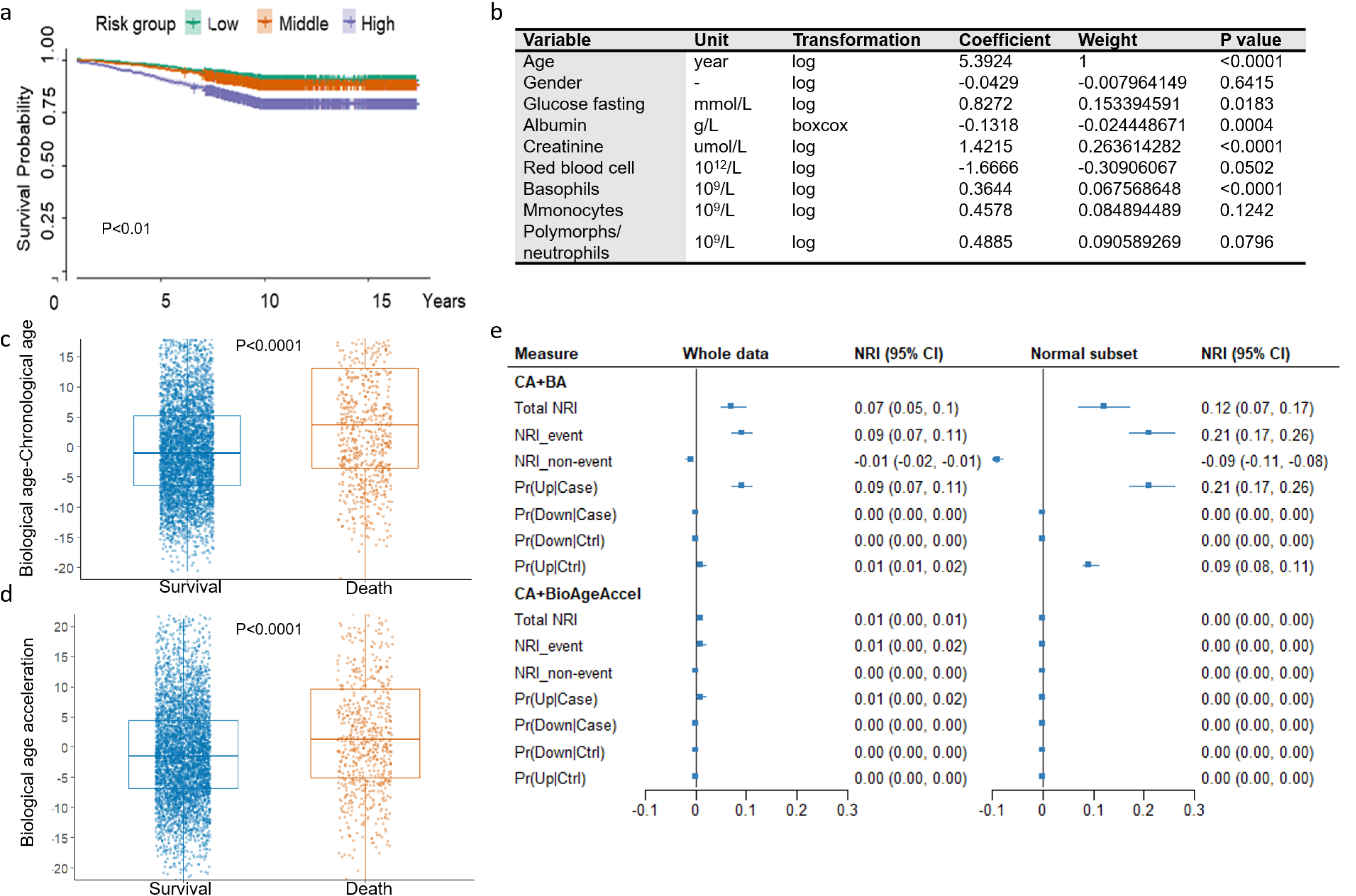
Evaluation of AQ in predicting mortality risk. (a) Kaplan–Meier survival curves for all-cause mortality in low-, middle-, and high-risk groups. Individuals with higher risk score exhibited significantly lower survival probabilities over follow-up (P<0.01). (b) Risk score system based on Cox model fitted to all-cause mortality. (c) Distribution of AQ difference with chronological age among survivors and deaths during follow-up. Survivals exhibited significantly lower AQ acceleration than deaths (P<0.0001). (d) Comparison of AQ acceleration between survival and deaths. Survivals demonstrated lower AQ acceleration (P<0.0001). Each dot corresponds to an individual participant. Blue dots represent survivals; orange dots represent deaths.

**Table 1.**
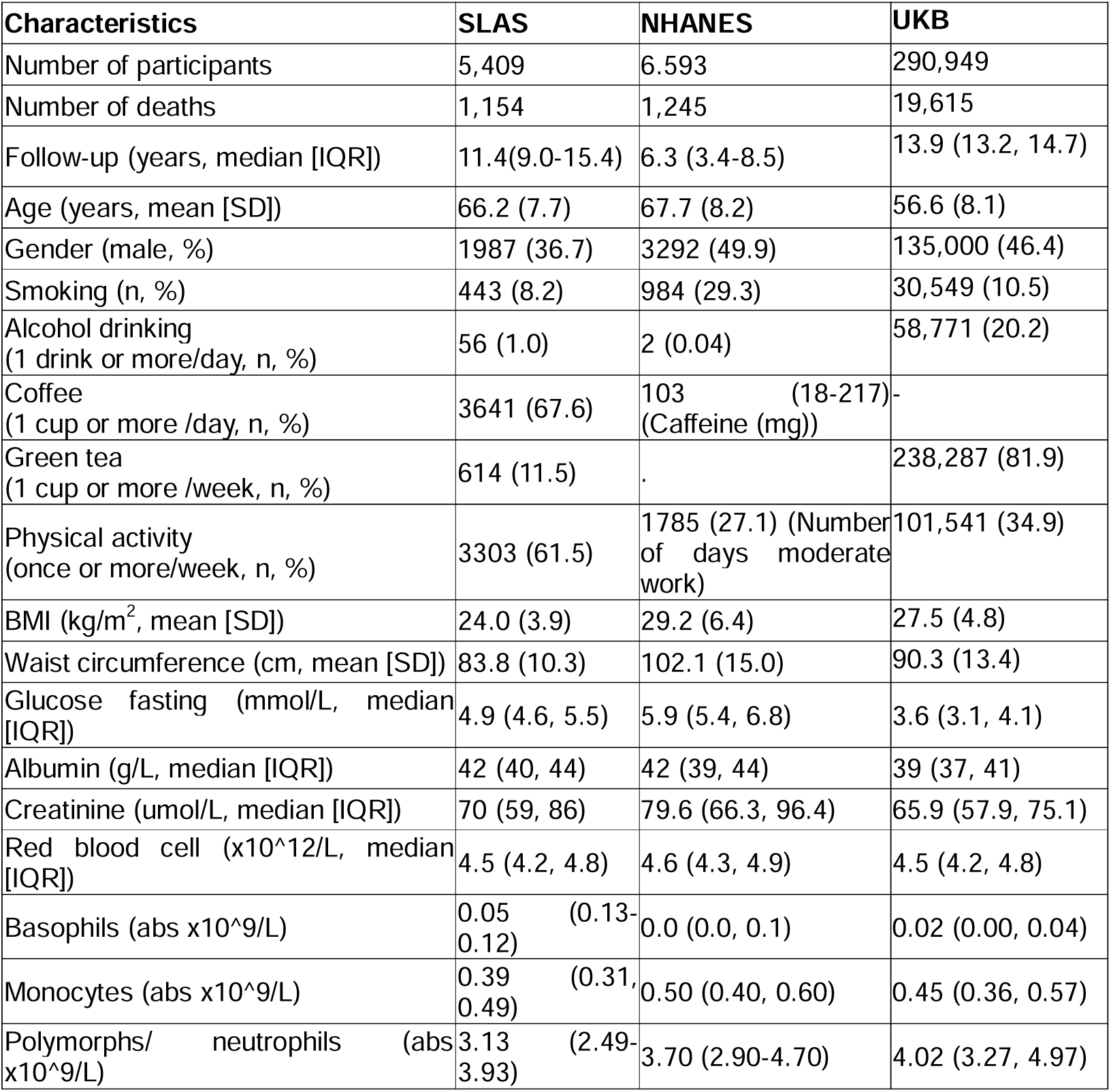
Baseline characteristics of the study participants from SLAS, NHANES and UKB.

**Table 2.** Overall and cause-specific mortality validation for the biological age in SLAS, NHANES, and UKB.

| Cause of death | SLAS |  |  |  |  | NHANES |  |  |  |  | UKB |  |  |  |  |
| --- | --- | --- | --- | --- | --- | --- | --- | --- | --- | --- | --- | --- | --- | --- | --- |
|  | Number of cases | BA (years, Median [IQR]) | HR | P value<sup>a</sup> | C-index | Number of cases | BA (years, Median [IQR]) | HR | P value<sup>a</sup> | C-index | Number of cases | BA (years, Median [IQR]) | HR | P value<sup>a</sup> | C-index |
| **All-cause** | 732 | 71.8 (60.0, 87.3) | 1.03 | <0.0001 | 0.629 | 1245 | 65.4 (53.5, 79.8) | 1.03 | <0.0001 | 0.721 | 19615 | 64.93[57.81,72.00] | 1.05 | <0.0001 | 0.706 |
| **Cancer** | 357 | 67.8 (59.2, 79.1) | 1.02 | <0.0001 | 0.580 | 295 | 56.6 (42.0, 77.8) | 1.02 | <0.0001 | 0.647 | 9614 | 63.80[56.85,70.48] | 1.05 | <0.0001 | 0.687 |
| **Cardiovascular** | 342 | 73.4 (61.5, 87.2) | 1.04 | <0.0001 | 0.672 | 338 | 67.7 (56.0, 81.6) | 1.04 | <0.0001 | 0.746 | 4086 | 66.51[59.32,73.99] | 1.05 | <0.0001 | 0.745 |
| **Respiratory diseases** | 211 | 73.7 (60.8, 87.1) | 1.03 | <0.0001 | 0.641 | 63 | 75.6 (63.9, 90.6) | 1.03 | <0.0001 | 0.707 | 1454 | 67.53[60.59,75.20] | 1.06 | <0.0001 | 0.777 |
| **Diabetes** | 10 | 77.2 (67.6, 114.1) | 1.06 | <0.0001 | 0.778 | 37 | 83.0 (72.4, 96.6) | 1.04 | <0.0001 | 0.789 | 161 | 74.01[64.47,81.78] | 1.06 | <0.0001 | 0.823 |
| **Alzheimer's** | 0 | - | - | - | - | 42 | 73.2 (65.8, 83.3) | 1.03 | <0.0001 | 0.727 | 285 | 65.53[60.05,71.86] | 1.05 | <0.0001 | 0.764 |

*a, P value for biological age*

We first considered the association of lifestyle factors and body weight in relation to all-cause and cause-specific mortality. Across SLAS, NHANES and UKB cohorts, smoking was consistently associated with higher all-cause mortality (SLAS HR 1.90, 95% CI 1.48–2.48; NHANES 2.13, 1.81–2.48; UKB 1.87, 1.82–1.91), with stronger associations for cardiovascular and respiratory mortality in UKB (**Fig. 2a-c, Extended Tables** 2-4). BMI was positively associated with all-cause mortality in SLAS and UKB (SLAS HR 1.03, 95% CI 1.01–1.05; UKB HR 1.02, 95% CI 1.01– 1.03), whereas no significant association was observed in NHANES (HR 1.00, 95% CI 0.98–1.01). In contrast, green tea intake was associated with lower all-cause mortality in SLAS (trend HR 0.86, 0.77–0.97) and UKB (e.g., 1–2 cups/day HR 0.85, 0.78–0.92), with additional reductions in cancer mortality in UKB. Physical activity showed modest protective associations with all-cause mortality, borderline effect in NHANES (HR 0.83, 0.66–1.04) and significant decreased risk in UKB (HR 0.91, 0.83–1.00), but not significant effect in SLAS. Plant food intake and coffee consumption showed no association, while alcohol intake exhibited heterogeneous inverse associations in UKB but not in SLAS and NHANES. When adjusted for waist circumference (WC), the associations between WC and all-cause mortality were minimal: SLAS HR 1.00 (95% CI 1.00–1.01), NHANES HR 1.00 (95% CI 1.00–1.01), and UKB HR 1.01 (95% CI 1.01–1.01); effects of various lifestyle factors also remained unchanged (results not shown). Overall, smoking was the most consistent risk factor, whereas green tea, and physical activity were protective but cohort-dependent.

**Figure 2.**
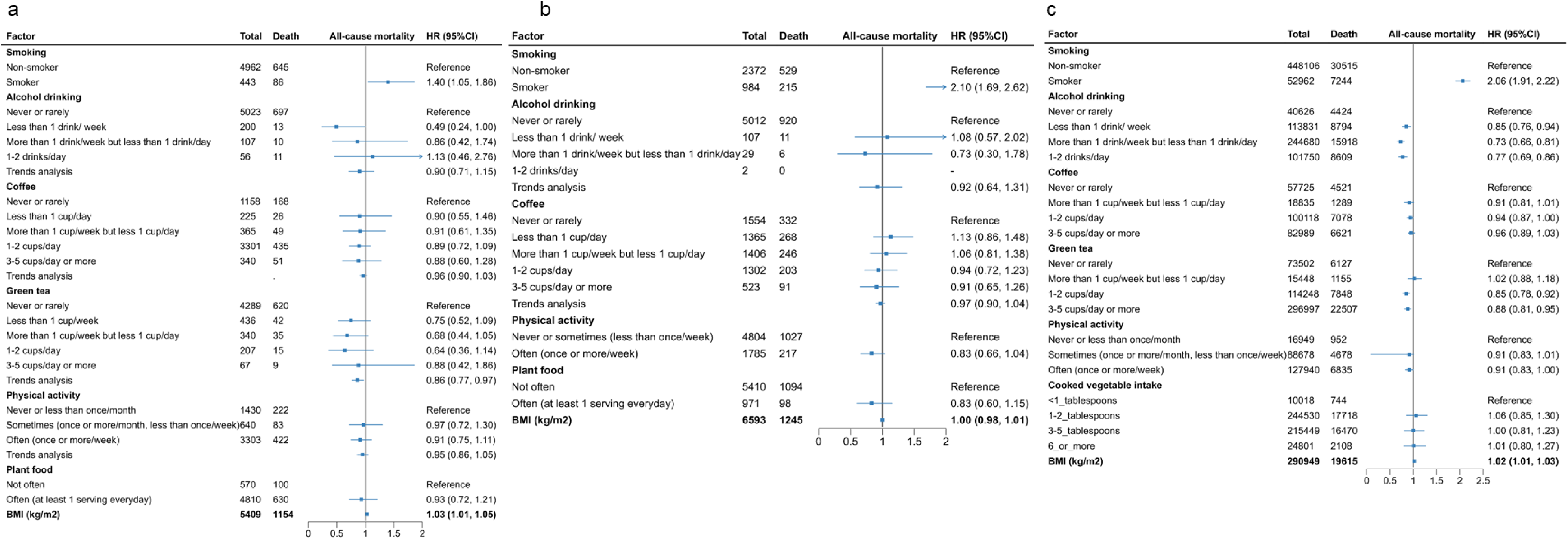
Association of lifestyle factors with all-cause mortality across three cohorts. (a) Singapore Longitudinal Ageing Studies (SLAS). (b) National Health and Nutrition Examination Survey (NHANES). (c) UK Biobank (UKB). Hazard ratios (HRs) and 95% confidence intervals (CIs) were estimated using multivariable models adjusted for age, sex, body mass index, and all listed lifestyle factors. Blue squares represent the estimated HRs, with error bars representing the 95% CIs. Vertical lines are reference line with HR = 1.0.

While the mechanisms linking positive lifestyle factors and body weight to better health outcomes are not well-defined, these can be plausibly attributed to slowing down a multi-domain biological ageing process ^11^. In SLAS, smoking was strongly associated with higher odds of AQ acceleration (OR 1.90, 95% CI 1.48–2.48; p<0.0001), whereas green tea intake, physical activity, and plant-based diet were consistently associated with lower odds (**Table 3**). BMI and WC demonstrated increased risk of biological age acceleration in SLAS (BMI: OR 1.07, 95% CI 1.05–1.09, p<0.0001; WC: OR 1.03, 95% CI 1.02–1.04, p<0.0001). Notably, green tea showed an inverse dose–response association (p for trend = 0.0007). Regular physical activity (≥ once per week) was associated with substantially reduced odds (OR 0.71, 95% CI 0.60–0.83; p <0.0001), and frequent plant food intake conferred similar protective effect (OR 0.59, 95% CI 0.50–0.72; p = 0.0126). Similar analyses were pursued across NHANES and UKB cohorts; smoking remained consistently associated with increased odds while higher plant food intake, green tea and regular physical activity with lower odds of AQ acceleration in UKB, but not significant in NHANES (**Table 3**). Both lifestyle factors and body weight were associated with AQ acceleration. These remained after accounting for baseline metabolic disease, including diabetes, hypertension and hypercholesterolemia.

**Table 3.**
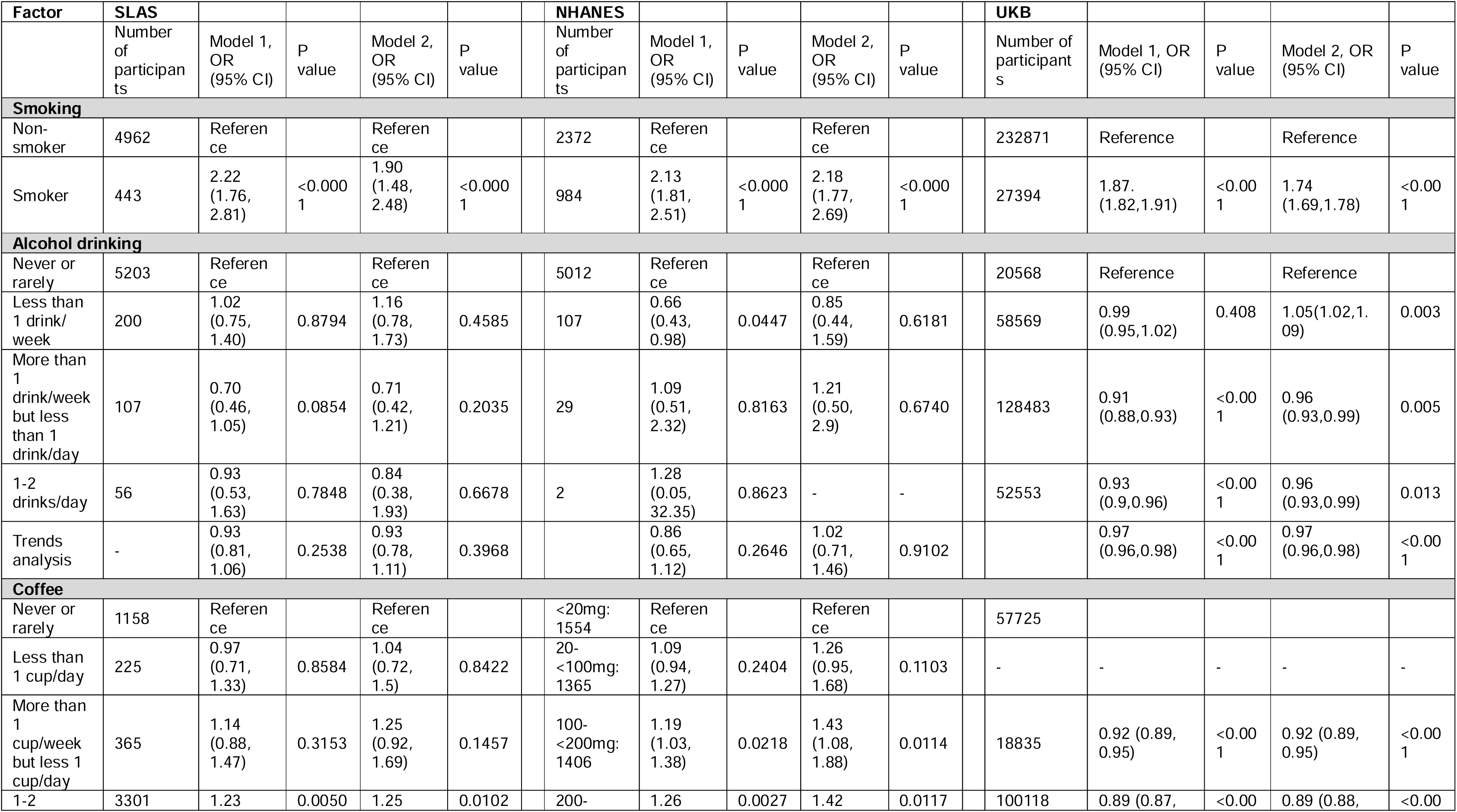
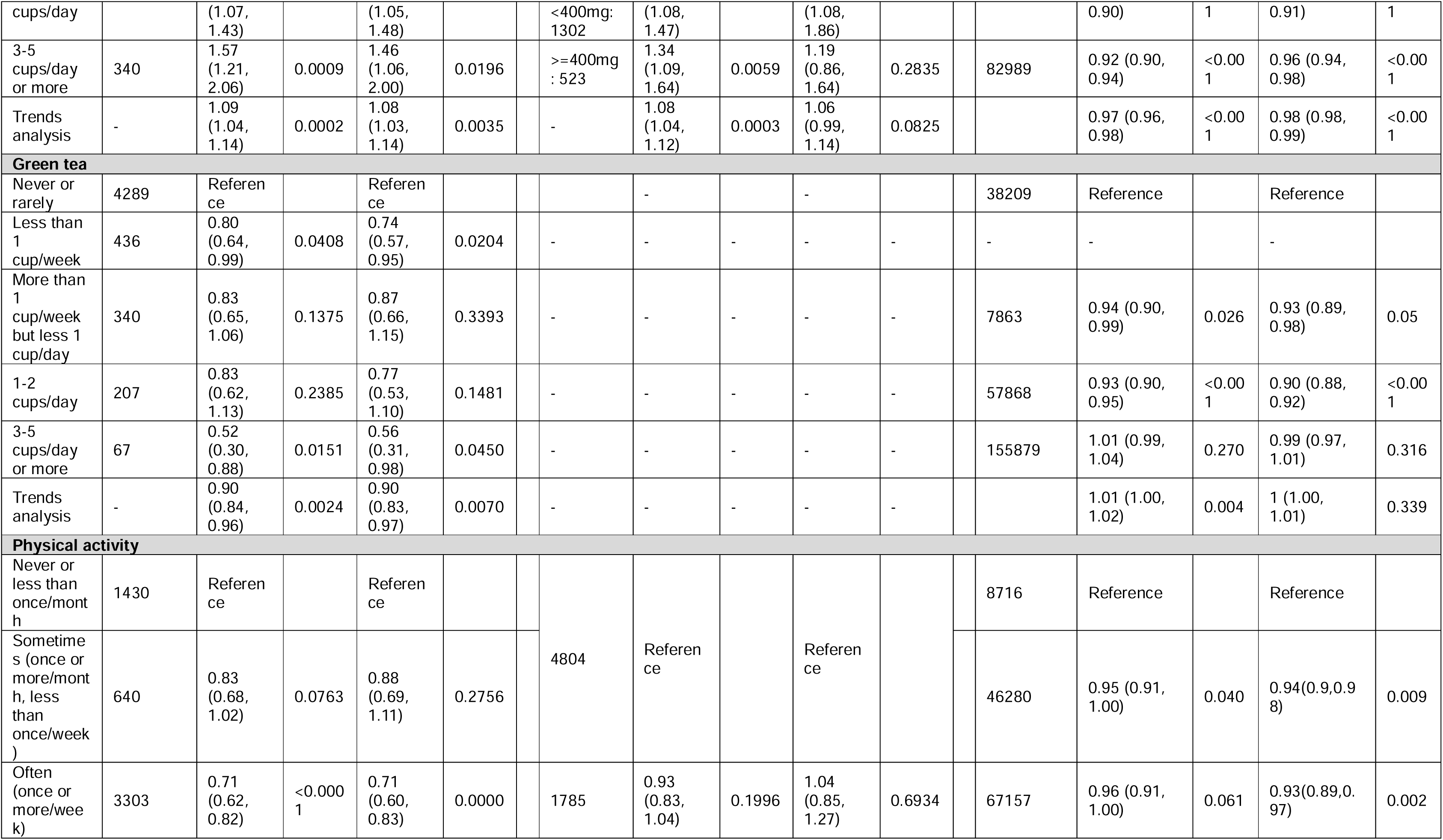
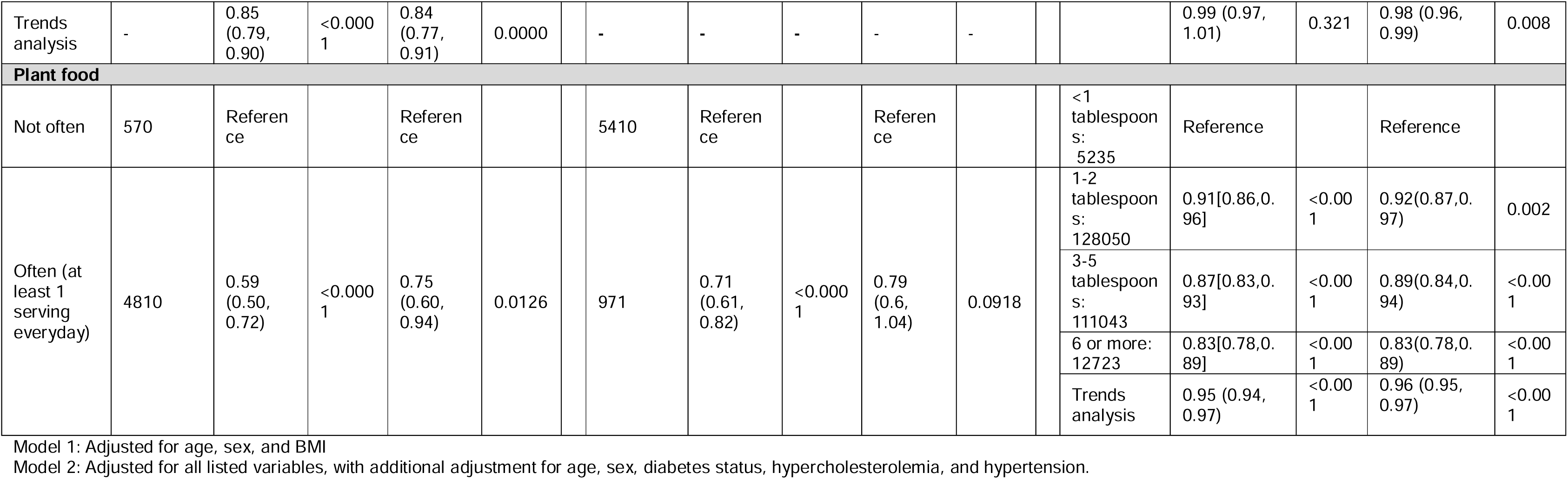
Association of lifestyle factors with biological age acceleration in SLAS, NHANES, and UKB.

We then sought to assess the extent to which AQ mediates the associations between lifestyle factors, body weight and health outcomes. Results support a partial mediating role of AQ in all-cause mortality across both SLAS and NHANES cohorts (**Fig. 3a-b**). The negative effect of smoking on AQ was significant in both SLAS (direct HR 1.35, 95% CI 1.05–1.73; indirect HR 1.10, 1.07–1.14) and NHANES (direct HR 1.66, 1.36–2.01; indirect HR 1.13, 1.09–1.17), with approximately 28% and 25% of the excess risk mediated, respectively. Green tea consumption (SLAS only) was associated with lower all-cause mortality (direct HR 0.88, 0.79–0.98; indirect HR 0.99, 0.98–1.00), with minimal mediation through AQ. For physical activity, a protective effect was observed in NHANES (HR 0.88, 0.79–0.97) but not in SLAS, and no significant mediation was evident. Plant food intake showed significant protective effects through AQ, as observed in both cohorts (SLAS: HR 0.97, 0.94–0.99; NHANES: HR 0.93, 0.89–0.97). This accounted for 18% (SE 14%) and 27% (SE 20%) of the associations, respectively, although direct effects were not statistically significant. Altogether, these findings highlight biological ageing as an important pathway potentially linking multiple lifestyle factors and body weight with mortality, although residual confounding and the observational design can limit causal interpretation. Associations were directionally consistent after additional adjustment for diabetes, hypertension, and lipid status, supporting robustness to cardiometabolic confounding.

**Figure 3.**
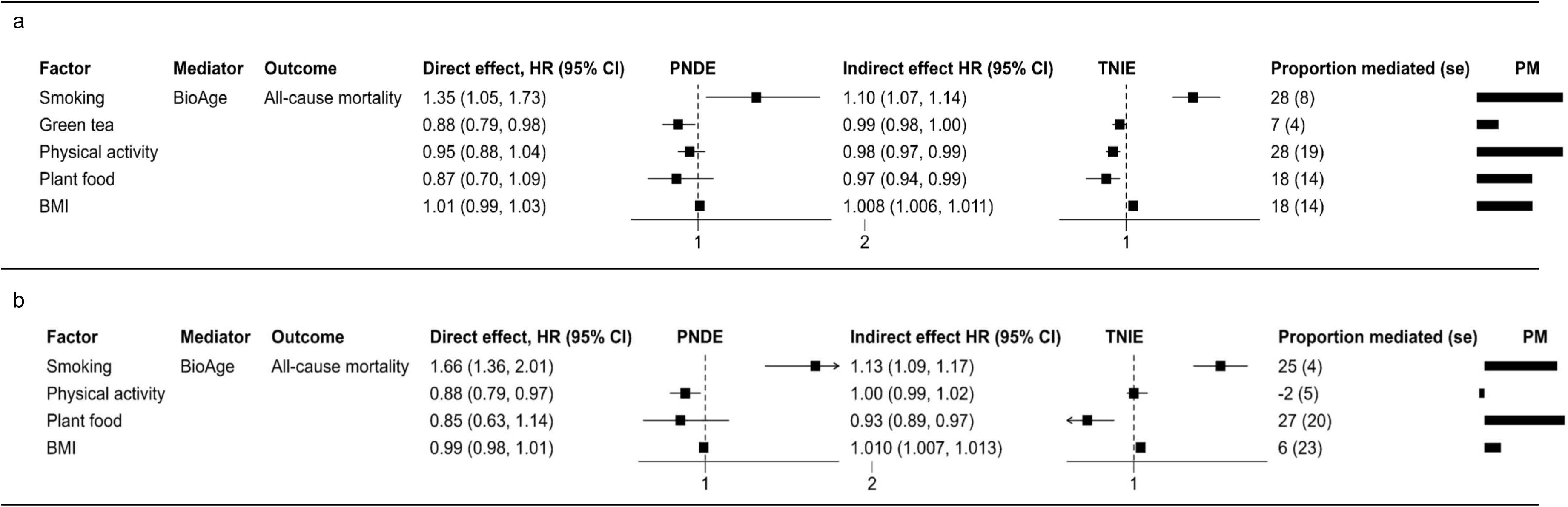
Mediating analyses of AQ acceleration in the associations between lifestyle factors and all-cause mortality. (a) Singapore Longitudinal Ageing Studies (SLAS). (b) National Health and Nutrition Examination Survey (NHANES). (c) UK Biobank (UKB). The squares represent the estimated HRs; error bars represent the 95% confidence intervals (CIs). The vertical lines represent reference line with HR = 1.0. The bars indicate the proportion of effects mediated by AQ acceleration.

To further investigate whether modifiable exposures are associated with AQ, 10-year all-cause mortality, and biological subsystems, we evaluated BMI, WC, and six lifestyle exposures (physical activity, green tea, coffee, smoking, alcohol, and plant food) in relation to AQ difference, mortality, and five AQ-derived domain measures. These domains include metabolic, nutrition/synthesis, clearance, circulatory, and inflammatory domains based on known biological association between candidate variables and organ function (see Methods). In the primary analysis, BMI, WC, and smoking showed the strongest associations with older AQ (**Fig. 4a-b**). Compared with normal BMI, obesity class II was associated with a 4.17-year higher AQ difference (95% CI 3.063 to 5.283). Substantially increased WC was associated with a 3.29-year higher AQ difference relative to normal WC (95% CI 2.527 to 4.056). Current smoking was associated with a 3.55-year higher AQ difference compared with non-smoking (95% CI 2.528 to 4.576). Physical activity and plant food were associated with lower AQ difference after multiplicity correction (**Fig. 4a-b**). Frequent physical activity was associated with a 1.25-year lower AQ difference relative to never or less-than-monthly activity (95% CI −1.856 to −0.636). Plant food intake was associated with a 1.02-year lower AQ difference compared with no plant food intake (95% CI −1.875 to −0.160). Green tea, alcohol, and coffee did not meet the false-discovery threshold for AQ difference in the primary results table. For 10-year all-cause mortality, the strongest adjusted evidence was again observed for BMI, WC, and smoking (**Fig. 4a**).

**Figure 4.**
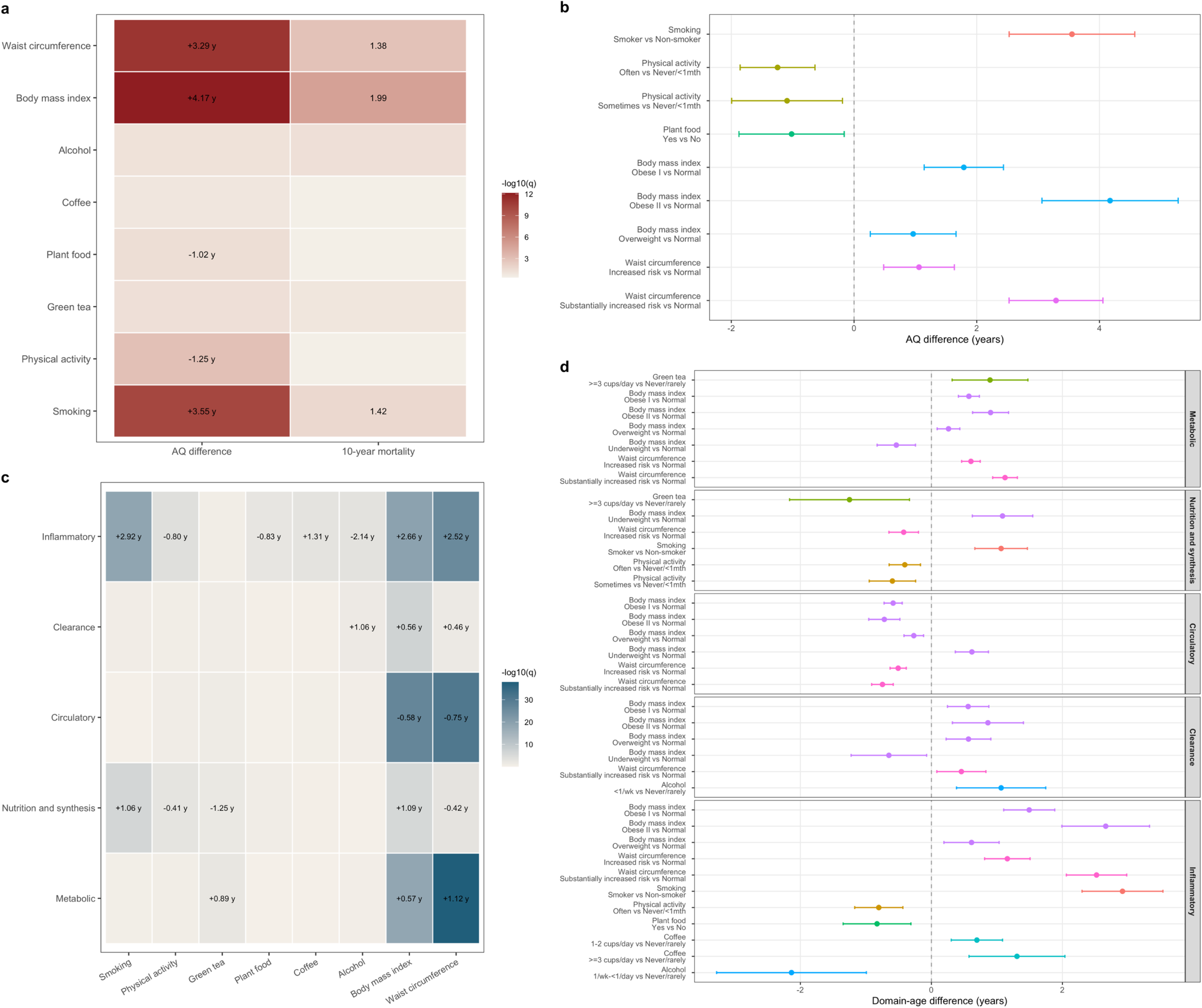
Primary and domain-specific associations of modifiable exposures with AQ measures and mortality. (a) Heat map of overall associations for the preferred primary model of each exposure across AQ difference and 10-year mortality. Tile colour indicates -log10(q); labels show representative effect estimates for associations with q < 0.05 (years for AQ difference and hazard ratios for mortality). (b) Forest plot of contrast estimates for AQ difference for exposure-outcome pairs with family-level q < 0.05. Points indicate effect estimates in years and horizontal lines indicate 95% confidence intervals. (c) Heat map of overall associations for domain-specific age measures. Tile colour indicates -log10(q); labels show representative effect estimates in years for associations with q < 0.05. (d) Forest plots stratified by age domain, showing all contrast estimates with detail-level q < 0.05 for exposure-domain pairs with family-level q < 0.05. Points indicate effect estimates in years and horizontal lines indicate 95% confidence intervals. q values are Benjamini-Hochberg-adjusted P values.

Domain-specific analyses showed that exposure associations were not uniform across AQ components (**Fig. 4c-d**). BMI and WC showed broad and high-confidence domain associations. WC was most strongly associated with the metabolic (substantially increased risk vs normal: +1.12 years) and inflammatory domain (+2.52 years), with additional evidence for the clearance domain (+0.46 years). BMI showed a similar pattern for metabolic (+0.57 years for obese I vs normal), inflammatory (+2.66 years for obese II vs normal), and clearance (+0.56 years for obese I vs normal) domains. BMI also showed a nonlinear nutrition/synthesis signal, with underweight participants having higher nutrition/synthesis AQ difference than normal-BMI participants (+1.09 years).

Lifestyle factors involving smoking, physical activity, plant food, coffee, alcohol, and green tea showed more selective domain profiles (**Fig. 4c-d**). Smoking was strongly associated with higher inflammatory (+2.92 years) and nutrition/synthesis (+1.06 years) AQ difference, but not with metabolic, circulatory, or clearance domains after correction. Frequent physical activity was associated with lower inflammatory (−0.80 years) and nutrition/synthesis (−0.41 years) AQ differences. Plant food intake was associated primarily with lower inflammatory AQ difference (−0.83 years). Coffee showed a domain-specific association with higher inflammatory AQ difference, strongest for at least three cups per day (+1.31 years). Alcohol showed mixed component signals: moderate intake of 1/week to less than 1/day was associated with lower inflammatory AQ difference (−2.14 years), whereas less-than-weekly intake was associated with higher clearance AQ difference (+1.06 years). High green tea intake was associated with higher metabolic AQ difference (+0.89 years) but lower nutrition/synthesis AQ difference (−1.25 years). Smoking, physical activity, plant food, coffee, alcohol, and green tea did not show robust circulatory-domain associations after false-discovery correction. Overall, these domain analyses supports a heterogeneous exposure profile. Smoking and physical activity showed coherent inflammatory and nutrition/synthesis patterns, plant food was most clearly related to the inflammatory component, and BMI and WC had the broadest AQ-domain footprint. These results should be presented as mechanistic patterning of AQ component signals, not as direct causal evidence.

Sleep duration was evaluated separately in the SLAS sleep sub-cohort (n = 2,612). The strongest sleep-related signal was observed for the clearance domain (overall spline q = 1.14e-4; nonlinearity q = 0.00777) (**Fig. 5a)**. Across the well-supported sleep range of 4.0 to 9.0 hours, the fitted clearance-domain AQ difference varied by 1.135 years. The fitted curve was nonlinear but did not show a clear U-shaped pattern with an internal optimum. Instead, the lowest fitted value occurred at the lower boundary of the observed range, and clearance-domain AQ difference increased with longer sleep duration. This boundary pattern should be interpreted cautiously and should not be taken as evidence that very short sleep is beneficial for clearance. Metabolic and circulatory domains showed smaller nonlinear sleep associations. Metabolic AQ difference had an overall spline q value of 0.0113 and nonlinearity q value of 0.00777, with a fitted mid-90% range of 0.418 years and a fitted minimum at approximately 6.39 hours. Circulatory AQ difference had an overall spline q value of 0.0355 and nonlinearity q value of 0.0323, with a fitted range of 0.329 years and a fitted minimum at approximately 6.09 hours. These patterns are best described as shallow U-shaped associations, with higher fitted domain AQ difference at shorter and longer sleep durations and the highest fitted values near the upper end of the plotted sleep range. Inflammatory and nutrition/synthesis domains did not show statistically robust fitted-shape evidence after multiplicity correction.

**Figure 5.**
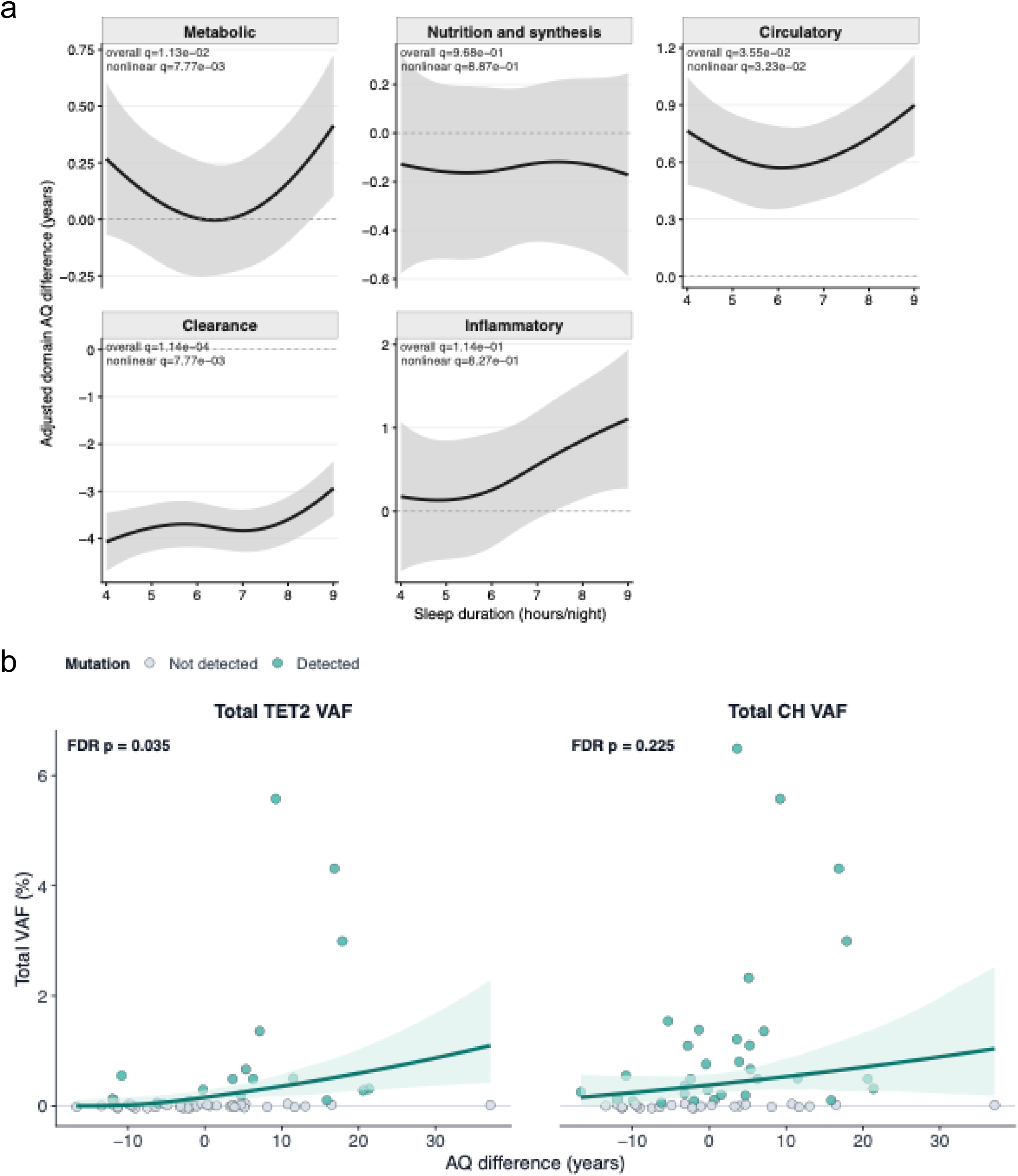
Sub-cohort Analyses for AQ-derived domain measures and TET2/CH mutation burden. (a) Sleep duration and AQ-derived domain measures. Adjusted natural-spline fits for sleep duration in the fixed sleep subcohort. Lines show fitted domain AQ difference across the middle 90% of observed sleep duration (4.0 to 9.0 hours/night), and shaded bands show pointwise 95% confidence intervals from the prediction standard error. Panel labels report Benjamini-Hochberg-adjusted overall spline q values and adjusted nonlinearity q values. (b) AQ difference and total TET2/CH mutation burden. Association between AQ difference (AQ minus chronological age) and total TET2 VAF or total CH VAF in 57 SLAS participants with available CH data. Points represent individual participants and are filled according to whether the corresponding mutation endpoint was detected. The fitted line and shaded band show age- and sex-adjusted marginal predictions from endpoint-specific log1p(VAF) linear regression models, back-transformed to the VAF percentage scale. FDR-adjusted p values are shown within each panel.

Lastly, since our study population encompassed individuals with ages corresponding to higher prevalence of clonal hematopoiesis (CH)-associated gene mutations ^12,13^, we also sought to investigate if associations existed between AQ and CH. CH gene mutation burden was analysed separately in the SLAS CH sub-cohort (n = 57). Higher AQ was associated most clearly with higher *TET2* mutation burden, while associations with overall CH burden were weaker and did not survive false discovery rate correction (**Fig. 5b**). In adjusted continuous burden models, each +1 year increase in AQ was associated with higher total *TET2* variant allele fraction (VAF) (beta = 0.0160) and max *TET2* VAF (beta = 0.0153). These findings show a higher baseline *TET2* burden in participants with higher AQ deviation, not as evidence of causality or longitudinal progression and suggest increased risk of CH with accelerated AQ. Given known links between CH and multiple diseases of aging, including cancer and cardiovascular disease, further investigation is warranted in a larger cohort.

## Discussion

In this study, we leveraged on routine laboratory biomarkers to predict mortality risk and estimate AQ. We demonstrate that individuals at high-risk of 10-year mortality exhibited AQ acceleration, with their AQ higher than chronological age. Consistently across all cohorts, smoking was associated with increased AQ acceleration while higher intake of plant-based diet conferred protective effect. We further showed that the associations between these lifestyle factors and mortality risk were, at least partly, mediated by their effects on biological ageing. Notably, at the domains level, where each domain reflects a biological component of the original AQ estimate model, we revealed mechanistic links through which lifestyle factors influence distinct biological domains, driving biological ageing and ultimately increasing mortality risk. For example, smoking was strongly associated with older inflammatory and nutrition/synthesis domains while frequent physical activity was associated with younger age of the same domains.

Smoking is a well-established risk factor for chronic disease and mortality ^14^, and our findings suggest that its detrimental effects may be mediated, at least in part, through accelerated biological ageing. Consistent with prior studies, smokers exhibit greater biological age acceleration compared to non-smokers ^15–19^, which has been positively linked to an increased risk of all-cause mortality ^20^. This supports the role of biological ageing as an intermediate phenotype mediating lifestyle pattern to adverse health outcomes. Importantly, large-scale studies reported that early age of smoking initiation appears to further amplify this effect, being associated with greater biological ageing acceleration and potentially mediated through heightened inflammation, oxidative stress and metabolic dysregulation ^15,16^.

Nutrition and diet are key determinants of health, particularly with the increasingly westernised lifestyle patterns characterised by higher intake of read and processed meat which are consistently linked to higher risks of chronic disease and mortality ^21–23^. Emerging evidence suggests that these associations may be partly driven by their impact on biological ageing, with proinflammatory and oxidative stress-inducing properties contributing to cumulative molecular and cellular damage ^24–26^. Diets that drive systemic inflammation and metabolic dysregulation may therefore accelerate biological ageing, increasing vulnerability to age-related disease and mortality. In contrast, plant-based diets preserve physiological function through anti-inflammatory and anti-oxidant effects of dietary fibers and polyphenols ^27^. These processes collectively decelerate biological ageing while enhancing resilience against disease and adverse health outcomes. However, it is also important to note that plant-based diets may also be unhealthful. While healthful diets involving whole grains, vegetables, and fruits, were associated with slower progression of biological ageing and reduced risks of chronic diseases and mortality, unhealthful diets appeared to accelerate biological ageing and were implicated with increased adverse outcomes^27–32^.

Frequent physical activity is a well-established protective factor against chronic diseases and mortality ^33,34^. Consistent epidemiological evidence linked low physical activity to an increased risk of non-communicable disease and higher mortality, while lifestyle improvement through increased physical activity is associated with improved health outcomes and longevity ^35–39^. These findings underscore its role as a key modifiable determinant of mortality risk. While the molecular mechanisms remains ill-defined, physical activity inhibited cancer cell proliferation, induced apoptosis, regulated cancer metabolism, and remodelled the immune microenvironment—processes implicated in tumor initiation and progression ^40^. In line with these insights, our findings that frequent physical activity is associated with younger inflammatory age are consistent with the concept of ‘inflamm-ageing’, a state of chronic, low-grade systemic inflammation that drives biological ageing ^41^. Frequent physical activity may mitigate this process by reducing systemic inflammatory burden and improving immune regulation ^42,43^.

Sleep is widely understood as essential for basic survival and many studies have investigated the relationship between inadequate sleep and a wide range of disorders, including hypertension, obesity, diabetes and various neurological and metabolic diseases ^44^. Our analysis of sleep duration showed associations with domain-specific aging, most robust signals associated with metabolic and circulatory AQ at internal optima between 6.1 and 6.4 hours. These findings align with prior evidence showing both short and excessively long sleep exacerbate physiological dysregulation and heighten metabolic risk ^45^. Notably, the clearance domain showed a different trajectory where longer sleep correlated with higher AQ. While the low-range boundary effects require cautious interpretation, these patterns may point to the possibility that systemic detoxification pathways are highly sensitive to sleep architecture and circadian disruption.

Moreover, since ageing is associated with gradual increase in the number of somatic mutations, the expansion of mutated hematopoietic stem cells may also lead to dysregulated biological systems. Expectedly, our analysis revealed a critical association between accelerated biological aging and the expansion of hematopoietic mutations. Participants with higher AQ exhibited a significantly greater burden of *TET2* mutations, which have been shown to drive systemic inflammation and independently elevate risks for cardiovascular disease and mortality ^46^. Our findings suggest that AQ deviation hint at a systemic environment permissive to clonal expansion or a state of reduced immunosurveillance as previously reported ^47^. With the small cohort size, further investigation is warranted to validate this finding.

Taken together, our findings highlight biological ageing as a plausible mechanistic link between modifiable lifestyle factors and long-term health outcomes. Rather than acting solely on independent risk factors, lifestyle behaviors may exert their effects on a shared ageing pathway that integrate multiple biological domains. Within this framework, we further identified domain-specific associations, indicating the lifestyle factors exert selective effects across biological domains. This has important implications for prevention and intervention strategies, suggesting that targeting biological age may represent a complementary approach to traditional intervention. Moreover, it highlights the potential of biological age, which can be estimated using routine laboratory data, as a dynamic biomarker for monitoring population health aimed to promote health longevity.

### Strengths and limitations

With routine laboratory data being underutilised, a notable strength of the present study is clinical data integration, whereby we used readily available laboratory data for analyses and model development, making it highly accessible across various clinical settings and ensuring ease of integration into current workflows. We showed that routine blood tests can offer an untapped, low-cost opportunity to build an evidence-based model for earlier and more accurate risk prediction, thereby reducing healthcare cost and improving screening accuracy. Additionally, our findings were validated in two independent large-scale cohorts from NHANES and UKB, supporting both scalability and generalizability across Asian, American, UK populations. This approach is adaptable to diverse populations, healthcare settings, and hospital systems, with the potential for re-training or transfer learning to ensure robust performance across contexts. However, there are also limitations to the study. Potential selection bias may affect generalisability of the findings. While our results were validated externally, the UKB recruitment process is volunteer-based, which could over-represent individuals who are more health conscious and differ from the general population. This is in contrast to the NHANS cohort that employs a strict selection criteria that randomly pick individuals from households to ensure accurate representation. Moreover, across all cohorts, lifestyle behavioural patterns were derived from self-reported data that may be subjected to recall bias and misclassification. Such subjective assessments may lead to measurement error and potential attenuation of observed associations. Future studies incorporating objective and longitudinal measurements, along with benchmarking of our AQ model against others, including Klemera and Doubal’s biological age (KDM-BA) ^48^, PhenoAge ^49^ and GrimAge ^50^, will be important to validate and strengthen the findings.

In conclusion, by analysing variations and interactions amongst biomarkers, our study provides an evidence-based framework for earlier and more accurate identification of high-risk individuals, offering a practical, inexpensive and easy-to-implement tool to inform preventive strategies across diverse populations.

## Method

### Study cohort

A predictive model for mortality was built using Singapore Longitudinal Aging Studies (SLAS) data and validated in the National Health and Nutrition Examination Survey (NHANES) and UK Biobank (UKB) datasets. Cohort descriptions are included below:

SLAS comprises of two independent multi-ethnic Singaporeans cohorts that were investigated for basic laboratory biomarkers (i.e. components of the blood cells, fasting glucose, cholesterol, proteins, etc.) and their relation to mortality. Participants aged 55 years and older, capable of self-ambulation were recruited in 2003 for SLAS-1 (median follow-up period of 17 years) and 2008 for SLAS-2 (median follow-up period of 12 years). Detailed description of cohort recruitment and data collection have been described in earlier papers ^51^.

NHANES is an on-going cross-sectional programme focused on public health of the American population using a complex, multi-stage probability design in 2-year cycle since 1999-2000 ^52^. It employs structured in-home interviews and standardised examinations at mobile examination centers to collect demographic, socioeconomic, dietary, and health-related data, along with medical and physiological measurements. Data from 22,754 participants aged 20 and above, collected between 1999-2018, were accessed. After data cleaning and exclusion of participants with missing values on predictors and biological aging, a total of 6,593 individuals were included as our validation cohort.

UKB is a multi-center prospective cohort with more than 500,000 participants, aged 37-73 years, recruited between 2006-2010 in the United Kingdom. Using a touch-screen questionnaire, it collects demographic, socioeconomic, environmental, lifestyle and health-related data. After data cleaning and exclusion of participants with missing values, a total of 290,949 individuals were included as our validation cohort.

### Measurements

At baseline, participants underwent comprehensive assessments through standardised questionnaire, interviews, clinical evaluations, and blood testing for lifestyle and clinical data collection. Anthropometric variables included age, BMI and WC. Lifestyle variables, such as smoking status, alcohol consumption, coffee and green tea intake, diet (plant food), and exercise frequency, were selected after systematic review of the questionnaires, which demonstrated associations between with mortality outcomes ^53^. Candidate laboratory variables eventually selected consisted of fasting glucose levels, albumin and creatinine levels, red blood cell count, and white blood cell count (i.e. basophils, monocytes, polymorphs/neutrophils).

### Mortality outcome

All-cause mortality and cause-specific mortality (cancer, cardiovascular disease, respiratory disease, and Alzheimer’s) were used as study outcomes. For the SLAS cohort, date and cause of death from baseline up to 31 December 2020 was determined using the participant’s unique National Registration Identity Card number for computerised record linkage with the National Death Registry through the National Disease Registry Office of the Ministry of Health. Cause of death was ascertained using International Classification of Diseases 10th Revision (ICD-10) codes.

NHANES mortality outcomes were obtained from the National Death Index of the National Center for Health Statistics (NCHS), while UKB mortality outcomes were derived from linked records provided by the NHS Information Centre (England and Wales) and the NHS Central Register (Scotland). Details have been described previously. ^52,54^.

### Statistical Analysis

The SLAS dataset contained 35 laboratory variables, of which 6 with more than 50% missing values were excluded. The remaining 29 variables were examined for collinearity. Variables with a Pearson correlation coefficient greater than 0.7 (P<0.05) with any other variables were excluded to reduce multicollinearity. This resulted in 18 retained laboratory variables, spanning three domains: complete blood count, blood lipids, and kidney function. Age and gender were included as fixed predictors yielding a total of 20 candidate predictors for model selection. Participants with missing values in any of the retained variables were excluded, resulting in a complete-case cohort of 5,409 individuals (88.9% of the total cohort) with complete data in the final analysis.

We checked the missing value, conducted a multiple imputation, and completed the sensitivity analyses with the imputed data. All candidate variables were assessed for normality using both visual inspection and formal statistical testing and appropriately transformed. For each variable, units, 5th-95th percentile range, median, and mean were reported.

### Model selection and development

A 10-fold cross-validated Cox regression with a Lasso penalty, repeated 10 times, was performed for model selection. The optimal penalty parameter (lambda) was selected from 100 candidate values, logarithmically spaced between 100 and 0.0001, to ensure model stability and robustness. An initial model incorporating 8 laboratory variables, along with age and gender as fixed covariates, was fitted. Predictors were subsequently evaluated using likelihood ratio tests, with those demonstrating P-values greater than 0.10 excluded. The final model retained 7 laboratory variables (P<0.10), in addition to age and gender, for the prediction of all-cause mortality. Validation was performed on the NHANES and UKB dataset, with appropriate adjustment.

### Biological age measurement

Biological age (age quotient, AQ) was estimated based on the selected Cox proportional hazards model fitted to all-cause mortality in the SLAS cohort. The validity of the estimated AQ was evaluated by examining its association with cause-specific mortality, including deaths due to cancer, cardiovascular, respiratory diseases, diabetes, and Alzheimer’s disease. Biomarkers used to derive AQ included chronological age, sex, complete blood count parameters, and routine biochemical measures collected at baseline. AQ acceleration was defined as the residuals from regressing estimated AQ on chronological age.

To examine whether AQ acceleration mediated the associations between lifestyle factors and mortality, causal mediation analyses was conducted within a Cox proportional hazards framework via the regmedint package to estimate the direct, indirect, and total effect of lifestyle exposures on the outcome, adjusting for age, sex, and BMI. Effect estimates were presented as the hazard ratios (HRs) with 95% confidence intervals (CIs) for mortality. The 95% CIs for direct and indirect effects, along with the proportions mediated, were estimated using simulation with 100,000 repeats.

### Domain-specific lifestyle associations

We further investigated the associations between lifestyle factors and domain-specific biomarker-linked aging signals. Biomarkers were centred to the reference population and aggregated into five domains: metabolic (glucose), nutrition/synthesis (albumin), circulatory (red blood cell), clearance (creatinine), and inflammatory (leukocyte). Outcomes were defined as domain age minus chronological age, with positive values indicating accelerated ageing. Lifestyle exposure models were mutually adjusted for nonlinear age, sex, education, ethnicity, and the remaining lifestyle exposures. BMI and WC were modelled using natural splines and adjusted for nonlinear age, sex, education, ethnicity, and all six lifestyle exposures. q values are Benjamini-Hochberg-adjusted P values. Sleep values outside 3 to 12 hours were excluded, and fitted curves were summarized over the middle 90% of observed sleep duration, corresponding to 4.0 to 9.0 hours. Each domain outcome was modelled using natural-spline linear regression for sleep duration, adjusted for nonlinear age, sex, education, ethnicity, physical activity, green tea, coffee, smoking, alcohol, and plant food intake. Overall spline P values tested the sleep spline term jointly, and nonlinearity P values compared the spline model with the corresponding linear sleep model.

In the 57-sample SLAS baseline subset, we assessed cross-sectional associations between AQ difference and clonal hematopoiesis (CH) mutation burden. AQ difference was defined as AQ minus chronological age. CH mutation calls were summarized at the participant level as CH and TET2 mutation burden, including maximum variant allele fraction (VAF) and total VAF. Samples without detected CH mutations were assigned zero mutation burden. Adjusted continuous burden models used linear regression with log1p(burden) as the outcome and included AQ difference, chronological age, and sex as covariates. False discovery rate (FDR) correction was applied across the tested continuous burden outcomes.

## Data availability

NHANES data are publicly available from the National Center for Health Statistics at https://www.cdc.gov/nchs/nhanes/index.htm. UK Biobank data are protected and not publicly available due to data privacy regulations; they are accessible to approved researchers for health-related research in the public interest through the UK Biobank Access Management System.

## Code availability

No customized code was developed in this study. Code used for data analysis will be made available from the corresponding author upon reasonable request.

