## Extended Tables for "A Blood-Based Biological Age Model Derived from Routine Laboratory Biomarkers in the Singapore Longitudinal Ageing Study"

**Extended Table 1. Mortality validation for AQ in NHANES**

| **Cause of death** |  |  |  |  |  |
| --- | --- | --- | --- | --- | --- |
|  | Number of cases | AQ (Median [IQR]) | HR | P value | C- index |
| All-cause | 1245 | 65.4 (53.5, 79.8) | 1.03 | <0.0001 | 0.721 |
| Cancer | 295 | 56.6 (42.0, 77.8) | 1.02 | <0.0001 | 0.647 |
| Cardiovascular | 338 | 67.7 (56.0, 81.6) | 1.04 | <0.0001 | 0.746 |
| Respiratory  diseases | 63 | 75.6 (63.9, 90.6) | 1.03 | <0.0001 | 0.707 |
| Diabetes | 37 | 83.0 (72.4, 96.6) | 1.04 | <0.0001 | 0.789 |
| Alzheimer’s | 42 | 73.2 (65.8, 83.3) | 1.03 | <0.0001 | 0.727 |

*Abbreviation: Age Quotient (AQ)*

**Extended Table 2. Association of lifestyle factors with cause-specific mortalities in SLAS**

| **Factor** | **Total number** | **Cancer mortality** | |  | **Cardiovascular diseases mortality** | |  | **Respiratory diseases mortality** | |  | **Diabetes mortality** | |
| --- | --- | --- | --- | --- | --- | --- | --- | --- | --- | --- | --- | --- |
|  |  | Death | HR (95% CI) |  | Death | HR (95% CI) |  | Death | HR (95% CI) |  | Death | HR (95% CI) |
| **Smoking** | | | | | | | | | | | | |
| Non-smoker | 4962 | 316 | Reference |  | 311 | Reference |  | 183 | Reference |  | 9 | Reference |
| Smoker | 443 | 41 | 1.30 (0.84, 2.03) |  | 31 | 1.23 (0.77, 1.97) |  | 27 | 2.24 (1.37, 3.66) |  | 1 | 4.55 (0.33, 63.53) |
| Alcohol drinking |  |  |  |  |  |  |  |  |  |  |  |  |
| Never or rarely | 5023 | 326 | Reference |  | 326 | Reference |  | 196 | Reference |  | 10 | Reference |
| Less than 1 drink/ week | 200 | 13 | 0.69 (0.32, 1.50) |  | 8 | 0.34 (0.11, 1.09) |  | 7 | 0.57 (0.21, 1.58) |  | 0 | - |
| More than 1 drink/week but less than 1 drink/day | 107 | 6 | 1.02 (0.42, 2.52) |  | 2 | 0.21 (0.03, 1.50) |  | 5 | 0.53 (0.13, 2.17) |  | 0 | - |
| 1-2 drinks/day | 56 | 11 | 2.29 (0.92, 5.68) |  | 5 | 1.06 (0.26, 4.34) |  | 1 | - |  | 0 | - |
| Trends analysis |  |  | 1.09 (0.84, 1.44) |  |  | 0.65 (0.41, 1.05) |  |  | 0.60 (0.34, 1.05) |  |  | - |
| **Coffee** | | | | | | | | | | | | |
| Never or rarely | 1158 | 79 | Reference |  | 67 | Reference |  | 40 | Reference |  | 1 | Reference |
| Less than 1 cup/day | 225 | 13 | 1.17 (0.62, 2.18) |  | 11 | 0.79 (0.36, 1.75) |  | 10 | 0.67 (0.24, 1.90) |  | 0 | - |
| More than 1 cup/week but less 1 cup/day | 365 | 29 | 1.03 (0.61, 1.74) |  | 19 | 0.93 (0.50, 1.72) |  | 12 | 0.72 (0.32, 1.64) |  | 2 | - |
| 1-2 cups/day | 3301 | 201 | 0.80 (0.59, 1.10) |  | 220 | 1.06 (0.77, 1.46) |  | 135 | 0.96 (0.65, 1.43) |  | 7 | - |
| 3-5 cups/day or more | 340 | 34 | 1.32 (0.8, 2.19) |  | 25 | 0.95 (0.52, 1.73) |  | 12 | 0.63 (0.29, 1.37) |  | 0 | - |
| Trends analysis |  |  | 0.96 (0.87, 1.06) |  |  | 1.02 (0.92, 1.12) |  |  | 0.98 (0.86, 1.10) |  |  | 2.19 (0.57, 8.47) |
| **Green tea** | | | | | | | | | | | | |
| Never or rarely | 4289 | 282 | Reference |  | 288 | Reference |  | 178 | Reference |  | 9 | Reference |
| Less than 1 cup/week | 436 | 26 | 0.92 (0.57, 1.50) |  | 21 | 0.70 (0.39, 1.26) |  | 14 | 0.76 (0.39, 1.51) |  | 0 | - |
| More than 1 cup/week but less 1 cup/day | 340 | 26 | 0.99 (0.59, 1.65) |  | 13 | 0.59 (0.30, 1.15) |  | 10 | 0.58 (0.26, 1.33) |  | 0 | - |
| 1-2 cups/day | 207 | 11 | 0.78 (0.39, 1.53) |  | 10 | 0.87 (0.43, 1.79) |  | 7 | 0.65 (0.24, 1.78) |  | 0 | - |
| 3-5 cups/day or more | 67 | 4 | 1.21 (0.45, 3.30) |  | 7 | 1.22 (0.45, 3.31) |  | 0 | - |  | 0 | - |
| Trends analysis |  |  | 0.97 (0.83, 1.12) |  |  | 0.91 (0.77, 1.07) |  |  | 0.77 (0.60, 0.99) |  |  | - |
| **Physical activity** | | | | | | | | | | | | |
| Never or less than once/month | 1430 | 108 | Reference |  | 112 | Reference |  | 55 | Reference |  | 3 | Reference |
| Sometimes (once or more/month, less than once/week) | 640 | 40 | 0.84 (0.55, 1.30) |  | 35 | 0.95 (0.61, 1.47) |  | 30 | 1.34 (0.78, 2.29) |  | 1 | - |
| Often (once or more/week) | 3303 | 207 | 0.81 (0.61, 1.09) |  | 194 | 0.86 (0.64, 1.16) |  | 125 | 1.13 (0.76, 1.66) |  | 6 | 0.82 (0.12, 5.50) |
| Trends analysis |  |  | 0.90 (0.78, 1.04) |  |  | 0.92 (0.80, 1.07) |  |  | 1.04 (0.86, 1.26) |  |  | 0.92 (0.34, 2.52) |
| **Plant food** | | | | | | | | | | | | |
| Not often | 570 | 39 | Reference |  | 39 | Reference |  | 34 | Reference |  | 1 | Reference |
| Often (at least 1 serving everyday) | 4810 | 317 | 1.00 (0.66, 1.51) |  | 303 | 1.09 (0.72, 1.66) |  | 176 | 0.70 (0.44, 1.10) |  | 9 | - |
| **BMI (kg/m^2^)** | 5409 | 1154 | 1.03 (1.00, 1.07) |  |  | 1.03 (1.00, 1.06) |  |  | 0.99 (0.95, 1.04) |  |  | 0.92 (0.71, 1.19) |

HRs (95% CIs) were derived from multivariable models including all listed variables, with additional adjustment for age, sex, diabetes status, hypercholesterolemia, and hypertension.

**Extended Table 3. Association of lifestyle factors with cause-specific mortalities in NHANES**

| **Factor** | **Total number** | **Cancer mortality** | |  | **Cardiovascular diseases mortality** | |  | **Respiratory diseases mortality** | |  | **Diabetes mortality** | |  | **Alzheimer’s** | |
| --- | --- | --- | --- | --- | --- | --- | --- | --- | --- | --- | --- | --- | --- | --- | --- |
|  |  | Death | HR (95% CI) |  | Death | HR (95% CI) |  | Death | HR (95% CI) |  | Death | HR (95% CI) |  | Death | HR (95% CI) |
| **Smoking** | | | | | | | | | | | | | | | |
| Non-smoker | 2372 | 141 | Reference |  | 124 | Reference |  | 39 | Reference |  | 18 | Reference |  | 16 | Reference |
| Smoker | 984 | 64 | 1.91 (1.29, 2.82) |  | 49 | 1.65 (1.00, 2.73) |  | 19 | 1.83 (0.89, 3.74) |  | 5 | 2.05 (0.57, 7.39) |  | 2 | 1.68 (0.17, 16.26) |
| Alcohol drinking |  |  |  |  |  |  |  |  |  |  |  |  |  |  |  |
| Never or rarely | 5012 | 233 | Reference |  | 247 | Reference |  | 49 | Reference |  | 26 | Reference |  | 29 | Reference |
| Less than 1 drink/ week | 107 | 3 | 0.89 (0.28, 2.84) |  | 2 | 0.60 (0.08, 4.33) |  | 2 | 2.31 (0.54, 9.83) |  | 0 | - |  | 0 | - |
| More than 1 drink/week but less than 1 drink/day | 29 | 2 | 1.00 (0.25, 4.09) |  | 2 | 0.82 (0.11, 5.92) |  | 0 | - |  | 1 | 10.70 (1.15, 99.58) |  | 0 | - |
| 1-2 drinks/day | 2 | 0 | (, ) |  | 0 | - |  | 0 | - |  | 0 | - |  | 0 | - |
| Trends analysis |  |  | 0.96 (0.52, 1.78) |  |  | 0.81 (0.32, 2.02) |  |  | 0.90 (0.29, 2.78) |  |  | 2.83 (0.90, 8.86) |  |  | - |
| **Coffee** | | | | | | | | | | | | | | | |
| <20mg | 1554 | 66 | Reference |  | 88 | Reference |  | 15 | Reference |  | 13 | Reference |  | 10 | Reference |
| 20- <100mg | 1365 | 60 | 1.53 (0.89, 2.61) |  | 80 | 1.09 (0.62, 1.89) |  | 8 | 0.56 (0.19, 1.62) |  | 6 | 2.26 (0.37, 13.92) |  | 8 | 4.71 (0.39, 56.78) |
| 100- <200mg | 1406 | 54 | 1.24 (0.72, 2.12) |  | 69 | 1.01 (0.59, 1.75) |  | 14 | 0.76 (0.31, 1.85) |  | 8 | 3.00 (0.53, 17.08) |  | 11 | 6.99 (0.77, 63.86) |
| 200- <400mg | 1302 | 66 | 1.27 (0.75, 2.15) |  | 44 | 0.60 (0.32, 1.11) |  | 10 | 0.57 (0.22, 1.50) |  | 5 | 1.40 (0.22, 8.87) |  | 8 | 5.43 (0.59, 50.3) |
| >=400mg | 523 | 27 | 1.32 (0.72, 2.43) |  | 25 | 0.84 (0.41, 1.72) |  | 11 | 1.61 (0.67, 3.90) |  | 4 | 1.88 (0.25, 13.86) |  | 0 | - |
| Trends analysis |  |  | 1.04 (0.91, 1.18) |  |  | 0.90 (0.78, 1.04) |  |  | 1.08 (0.86, 1.35) |  |  | 1.08 (0.73, 1.59) |  |  | 1.31 (0.82, 2.11) |
| **Physical activity** | | | | | | | | | | | | | | | |
| Never or less than once/month | 4804 | 236 | Reference |  | 289 | Reference |  | 51 | Reference |  | 33 | Reference |  | 40 | Reference |
| Sometimes (once or more/month, less than once/week) |  |  |  |  |  |  |  |  |  |  |  |  |  |  |  |
| Often (once or more/week) | 1785 | 59 | 0.92 (0.62, 1.39) |  | 48 | 0.57 (0.31, 1.02) |  | 12 | 0.92 (0.42, 2.04) |  | 4 | 0.26 (0.03, 2.01) |  | 2 | - |
| **Plant food** | | | | | | | | | | | | | | | |
| Not often | 5410 | 253 | Reference |  | 297 | Reference |  | 56 | Reference |  | 34 | Reference |  | 39 | Reference |
| Often (at least 1 serving everyday) | 971 | 28 | 0.98 (0.57, 1.69) |  | 25 | 0.69 (0.32, 1.51) |  | 4 | 0.79 (0.28, 2.24) |  | 2 | - |  | 1 | - |
| **BMI (kg/m^2^)** | 6593 | 1245 | 1.01 (0.98, 1.04) |  |  | 1.01 (0.98, 1.05) |  |  | 0.94 (0.89, 1.00) |  |  | 1.07 (0.99, 1.17) |  |  | 0.99 (0.87, 1.13) |

HRs (95% CIs) were derived from multivariable models including all listed variables, with additional adjustment for age, sex, diabetes status, hypercholesterolemia, and hypertension.

**Extended Table 4. Association of lifestyle factors with cause-specific mortalities in UK Biobank**

| **Factor** | **Total number** | **All-cause mortality** | | | | **Total number** | **Cancer mortality** | | | **Total number** | **Cardiovascular diseases mortality** | | **Total number** | **Respiratory diseases mortality** | | | **Total number** | | **Diabetes mortality** | | |
| --- | --- | --- | --- | --- | --- | --- | --- | --- | --- | --- | --- | --- | --- | --- | --- | --- | --- | --- | --- | --- | --- |
|  |  | Death | | HR (95% CI) | |  | Death | HR (95% CI) | |  | Death | HR (95% CI) |  | Death | | HR (95% CI) |  | | Death | HR (95% CI) | |
| **Smoking** | | | | | | | | | | | | | | | | | | | | | |
| Non-smoker | 232871 | 15857 | | 1.00 (Ref) | | 224875 | 7860 | Reference | | 220215 | 3201 | Reference | 218060 | 1046 | | Reference | 217154 | | 140 | Reference | |
| Smoker | 27394 | 3706 | | 2.06 (1.91-2.22) | | 25430 | 1742 | 2.03 (1.83-2.26) | | 24555 | 867 | 2.39 (2.03-2.83) | 24091 | 403 | | 2.67 (1.92-3.70) | 23708 | | 20 | 1.20 (0.36-3.99) | |
| **Alcohol** drinking |  |  | |  | |  |  |  | |  |  |  |  |  | |  |  | |  |  | |
| Never or rarely | 20568 | 2244 | | 1.00 (Ref) | | 19241 | 916 | Reference | | 18808 | 484 | Reference | 18551 | 227 | | Reference | 18356 | | 32 | Reference | |
| Less than 1 drink/ week | 58569 | 4557 | | 0.85 (0.76-0.94) | | 56248 | 2236 | 1.00 (0.86-1.16) | | 54950 | 938 | 0.74 (0.58-0.93) | 54372 | 360 | | 0.63 (0.42-0.95) | 54061 | | 49 | 0.92 (0.28-3.02) | |
| More than 1 drink/week but less than 1 drink/day | 128483 | 8358 | | 0.73 (0.66-0.81) | | 124376 | 4251 | 0.91 (0.79-1.05) | | 121860 | 1735 | 0.64 (0.52-0.79) | 120664 | 539 | | 0.46 (0.32-0.67) | 120178 | | 53 | 0.54 (0.16-1.76) | |
| 1-2 drinks/day | 52553 | 4398 | | 0.77 (0.69-0.86) | | 50344 | 2189 | 1.00 (0.86-1.16) | | 49071 | 916 | 0.65 (0.52-0.82) | 48478 | 323 | | 0.43 (0.28-0.66) | 48182 | | 27 | 1.22 (0.36-4.14) | |
| Trends analysis | Beta=-0.077, P < 0.001 | | | | | Beta=-0.004, P = 0.847 | | | | Beta=-0.115, P = 0.001 | | | Beta=-0.271, P < 0.001 | | | | Beta=0.035, P = 0.867 | | | | |
| **Coffee** | | | | | | | | | | | | | | | | | | | | | |
| Never or rarely | 57725 | 4521 | | Reference | | 55341 | 2137 | Reference | | 54188 | 984 | Reference | 53577 | 373 | | Reference | 53248 | | 44 | Reference | |
| More than 1 cup/week but less 1 cup/day | 18835 | 1289 | | 0.91 (0.81-1.01) | | 18170 | 624 | 0.85 (0.73-0.98) | | 17823 | 277 | 0.99 (0.78-1.27) | 17644 | 98 | | 1.31 (0.84-2.04) | 17562 | | 16 | 1.21 (0.37-3.94) | |
| 1-2 cups/day | 100118 | 7078 | | 0.94 (0.87-1.00) | | 96651 | 3611 | 0.92 (0.84-1.01) | | 94445 | 1405 | 0.89 (0.76-1.05) | 93505 | 465 | | 0.82 (0.60-1.13) | 93089 | | 49 | 0.53 (0.21-1.30) | |
| 3-5 cups/day or more | 82989 | 6621 | | 0.96 (0.89-1.03) | | 79577 | 3208 | 0.90 (0.81-1.00) | | 77765 | 1397 | 0.96 (0.81-1.13) | 76871 | 503 | | 1.13 (0.81-1.57) | 76419 | | 51 | 0.57 (0.22-1.47) | |
| Trends analysis | Beta=-0.013, P = 0.286 | | | | | Beta=-0.029, P = 0.083 | | | | Beta=-0.020, P = 0.482 | | | Beta=0.011, P = 0.848 | | | | Beta=-0.231, P = 0.139 | | | | |
| **Green tea** | | | | | | | | | | | | | | | | | | | | | |
| Never or rarely | 38209 | 3158 | | Reference | | 36534 | 1483 | Reference | | 35753 | 702 | Reference | 35294 | 243 | | Reference | 35092 | | 41 | Reference | |
| More than 1 cup/week but less 1 cup/day | 7863 | 589 | | 1.02 (0.88-1.18) | | 7576 | 302 | 0.98 (0.80-1.20) | | 7399 | 125 | 0.93 (0.66-1.32) | 7307 | 33 | | 0.84 (0.42-1.65) | 7276 | | 2 | 0.70 (0.08-5.91) | |
| 1-2 cups/day | 57868 | 3946 | | 0.85 (0.78-0.92) | | 55850 | 1928 | 0.83 (0.74-0.93) | | 54727 | 805 | 0.85 (0.71-1.03) | 54173 | 251 | | 0.64 (0.44-0.94) | 53956 | | 34 | 1.05 (0.37-2.96) | |
| 3-5 cups/day or more | 155879 | 11802 | | 0.88 (0.81-0.95) | | 149937 | 5859 | 0.87 (0.78-0.96) | | 146506 | 2429 | 0.88 (0.74-1.04) | 144988 | 911 | | 0.80 (0.58-1.11) | 144160 | | 83 | 0.68 (0.25-1.86) | |
| Trends analysis | Beta=-0.043, P < 0.001 | | | | | Beta=-0.043, P = 0.012 | | | | Beta=-0.040, P = 0.168 | | | Beta=-0.061, P = 0.283 | | | | Beta=-0.125, P = 0.440 | | | | |
| **Physical activity** | | | | | | | | | | | | | | | | | | | | | |
| Never or less than once/month | 8716 | 496 | | Reference | | 8481 | 261 | Reference | | 8312 | 92 | Reference | 8252 | 32 | | Reference | 8223 | | 3 | Reference | |
| Sometimes (once or more/month, less than once/week) | 46280 | 2425 | | 0.91 (0.83-1.01) | | 45214 | 1359 | 0.96 (0.84-1.09) | | 44284 | 429 | 0.88 (0.70-1.10) | 43974 | 119 | | 0.73 (0.48-1.09) | 43866 | | 11 | 0.65 (0.18-2.37) | |
| Often (once or more/week) | 67157 | 3559 | | 0.91 (0.83-1.00) | | 65489 | 1891 | 0.92 (0.81-1.05) | | 64350 | 752 | 1.04 (0.84-1.30) | 63772 | 174 | | 0.71 (0.47-1.05) | 63620 | | 22 | 0.99 (0.29-3.33) | |
| Trends analysis | Beta=-0.026, P = 0.202 | | | | | Beta=-0.042, P = 0.121 | | | | Beta=0.090, P = 0.053 | | | Beta=-0.116, P = 0.192 | | | | Beta=0.177, P = 0.535 | | | | |
| **Plant food** | | | | | | | | | | | | | | | | | | | | | |
| <1_tablespoons | 5235 | 385 | | Reference | | 5029 | 179 | Reference | | 4943 | 93 | Reference | 4884 | 34 | | Reference | 4854 | | 4 | Reference | |
| 1-2_tablespoons | 128050 | 9305 | | 1.06 (0.85-1.30) | | 123331 | 4586 | 0.99 (0.75-1.31) | | 120649 | 1904 | 1.00 (0.63-1.58) | 119435 | 690 | | 1.45 (0.46-4.57) | 118827 | | 82 | 0.69 (0.09-5.20) | |
| 3-5_tablespoons | 111043 | 8431 | | 1.00 (0.81-1.23) | | 106823 | 4210 | 0.92 (0.70-1.22) | | 104342 | 1730 | 0.94 (0.59-1.49) | 103208 | 596 | | 1.44 (0.46-4.54) | 102681 | | 69 | 0.47 (0.06-3.66) | |
| 6_or_more | 12723 | 1089 | | 1.01 (0.80-1.27) | | 12113 | 479 | 0.87 (0.64-1.20) | | 11893 | 259 | 0.95 (0.57-1.57) | 11720 | 86 | | 1.66 (0.49-5.61) | 11640 | | 6 | 0.26 (0.02-4.16) | |
| Trends analysis | Beta=-0.037, P = 0.073 | | | | | Beta=-0.065, P = 0.019 | | | | Beta=-0.043, P = 0.354 | | | Beta=0.045, P = 0.624 | | | | Beta=-0.411, P = 0.146 | | | | |
| BMI | 259736 | | 19426 | | 1.02 (1.01-1.03) | 249885 | 9574 | | 1.02 (1.01-1.03) | 244351 | 4041 | 1.04 (1.03-1.06) | 241743 | 1433 | 1.01 (0.98-1.05) | | 240465 | 155 | | | 0.99 (0.91-1.07) |

HRs (95% CIs) were derived from multivariable models including all listed variables, with additional adjustment for age, sex, diabetes status, hypercholesterolemia, and hypertension.
